# HbF/F-cell and the Phenotype of Sickle Cell Disease

**DOI:** 10.64898/2026.06.02.26354737

**Authors:** J. Lofters, A. Wilks, J. Lee, J. N. Hicks, E. S. Klings, M. H Steinberg

## Abstract

Fetal hemoglobin (HbF) prevents the polymerization of sickle hemoglobin (HbS). HbF, measured usually as a percent of total hemoglobin (%HbF), is inversely associated with the severity of sickle cell disease (SCD) but fails to capture the distribution of HbF concentrations within red blood cells (RBCs). The relative proportion of HbF and HbS within a RBC is reflected by the HbF:HbS ratio whereas HbF/F-cell quantifies the absolute amount of HbF/RBC. While correlated, HbF:HbS ratio and HbF/F-cell are not interchangeable. In the context of mean corpuscular hemoglobin (MCH), HbF/F-cell approximates whether sufficient HbF is present to inhibit HbS polymerization. We examined the association of mean HbF/F-cell with sub-phenotypes of sickle cell disease in three independent cohorts. Both %HbF and HbF/F-cell were significantly associated with multiple clinical and laboratory features of SCD; however, HbF/F-cell demonstrated stronger associations with clinical severity measures across cohorts. Higher HbF/F-cell was associated with fewer clinical events, reduced hemolysis, and mortality. Changes in HbF/F-cell after hydroxyurea treatment were associated with ~11-13% reduction in acute events in patients with <1 pg increase and >60% reduction with a >5 pg increase in HbF/F-cell. For each pg increase in HbF/F-cell there was ~6% reduction in the rate of acute events. As a surrogate for the distribution of HbF concentrations among F-cells, HbF/F-cell adds physiologically relevant insights that could guide prognosis and treatment.

**Key points:**

- HbF/F-cell reflects protective intracellular HbF:HbS stoichiometry.
- Higher HbF/F-cell demonstrated stronger associations with clinical severity than %HbF and predicts VOEs and ACS, reduced hemolysis, and mortality.
- Regardless of %HbF, as MCH decreases, the HbF:HbS ratio increases
- HbF/F-cell is a surrogate for the distribution of HbF concentrations among F-cells and could guide prognosis and treatment.

## Introduction

Fetal hemoglobin (HbF) prevents the polymerization of deoxy sickle hemoglobin (HbS). The HbS polymer is primarily responsible for the red blood cell (RBC) injury that triggers the pathophysiology of sickle cell disease (SCD).^1–3^ To a first approximation, %HbF is inversely associated with disease severity.^4–8^ The two clinically available HbF-related measurements, HbF and %F-cells, do not reflect distribution of HbF concentrations among RBCs (**Table S1**).^9, 10^ Sickle RBCs appear to be fully protected from HbS polymer-induced injury when the HbF:HbS ratio is ~0.30.This is exemplified by the phenotype of HbS-gene deletion hereditary persistence of HbF (HPFH) and by the “functional cure” after HbF-inducing gene therapy.^11–14^ In a sickle RBC with a normal mean corpuscular hemoglobin (MCH) of ~30 pg, ~10 pg of HbF is needed to achieve this ratio.

MCH increases when hydroxyurea (HU) treatment induces HbF.^15, 16^; the highest levels of HbF are associated with the highest MCH.^17^ In all in patients with the HbS/F genotypes (HbS homozygotes, HbS-β^0^ thalassemia), regardless of HU use, HbF is usually distributed unevenly (heterocellularly) among sickle RBCs. As MCH increases, more HbF/F-cell is required to maintain a HbS polymerization-inhibiting HbF:HbS ratio of ~0.30. Based on the contrast between the hematologic and clinical effects of HU treatment and gene therapy, we hypothesized that at a given %HbF, patients with lower MCH should have fewer disease complications because their RBCs have a more favorable HbF:HbS ratio. We calculated mean HbF/F-cell, using it as a surrogate for the distribution of HbF concentrations among F-cells, in order to examine its relationship with clinical outcomes of SCD. We propose that HbF/F-cell could serve as a widely available biomarker for SCD that might help guide patient care and clinical trial design.

## Methods

### Modelling MCH and HbF:HbS ratio

We first modelled MCH variation and its effect on HbF:HbS ratio, calculating the proportion of sickle RBCs with a fully protective HbF:HbS ratio of ~0:30, assuming each F-cell has a fixed mass of HbF (pg) corresponding to the %HbF measured at an MCH reference value of 30 pg. HbF:HbS ratio was calculated as: HbF:HbS = HbF mass (pg)/MCH − HbF mass (pg). We then considered a hypothetical RBC population whose HbF mass/cell follows a positively skewed distribution consistent with empirical measurements of F-cells, using a log-normal distribution to reflect this biological skew: HbF mass (pg) ~ Lognormal(*μ* = log(5), *σ* = 0.35), with values truncated at 0.1–22 pg reflecting physiologic bounds of HbF/F-cell and to avoid exceeding MCH at any simulated value. HbF/F-cell was computed as: HbF/F-cell = MCH × (% HbF) /100. HbF/F-cell represents the average mass of HbF per RBC. This distribution was held constant across all conditions to represent a fixed biological “input” at different MCH levels. MCH values of 25 pg and 35 pg approximating physiologic levels in the HbS/F only genotypes of SCD were modeled. The numerator was fixed for each simulated cell, while the denominator (HbS mass approximated as MCH − HbF mass) increased with MCH. A HbF:HbS ratio of ~0.30 was prespecified as the putative protective threshold.

### Cohorts

To examine the association of MCH with features of sickle cell disease, we examined data from of three groups of patients with HbS/F genotypes: 1. Cooperative Study of Sickle Cell Disease database (CSSCD, 1978-1988); 2. Multicenter Study of Hydroxyurea in Sickle Cell Disease database (MSH, 1992-1995); 3. Boston Medical Center registry (BMC, 2015-2025).^15,18^ The CSSCD was completed in the pre-HU era; patients in the MSH were treated with HU in a controlled clinical trial where treatment-related change in HbF (ΔHbF) could be measured; BMC patients represent current treatment paradigms and are included as a contemporary validation cohort (Table 1). In the MSH cohort, 150 patients were treated with HU, while 149 received placebo. Patients chronically transfused or transfused within 6 wks of data extraction were excluded from analysis. Definitions of clinical events and methods for laboratory testing were reported previously.^4,15,19–23^ The study was approved by the Institutional Review Board at the Boston University Medical Campus and Boston Medical Center.

**Table 1.** Cohort characteristics. Demographics of three cohorts; CSSCD, MSH and BMC. *HbF at baseline

| Cohort | N | Timeframe | Male (%) | Age | % on Hydrea | HbF% | HbF/F-cell (pg) | MCH (pg) |
| --- | --- | --- | --- | --- | --- | --- | --- | --- |
| **CSSCD** | 1742 | 1978-1988 | 49 | 22.8±12.4 | 0 | 6.2±6.0 | 1.56±1.91 | 28.9±3.2 |
| **MSH** | 150 | 1992-1995 | 49 | 30.2 ± 7.5 | 100 | *5.0±3.5 | 7.89 ± 6.67 | 32.72±4.6 |
| **BMC** | 46 | 2015-2025 | 39 | 40.7±7.9 | 76 | 11.6±8.8 | 4.2±3.8 | 33.0±5.4 |

### Data Analysis

We used standard statistical programming tools in R (with common packages) with widely used libraries for data handling, statistics, and visualization. Code development and verification were assisted using large language model-based coding support tools (ChatGPT, Open AI); all final analyses and outputs were independently validated in R/Python. Random-seed control was applied to ensure reproducibility. Outcomes such as acute vasoocclusive (VOE), were evaluated using ordinary least squares linear regression via the lm() function. Binary outcomes including acute chest syndrome (ACS), avascular necrosis (AVN), stroke, death, and priapism were modeled using multiple logistic regression implemented in glm (family = binomial). All models were adjusted for age, sex, and time in study; priapism analyses were adjusted for age and study time only. Model outputs were summarized as regression coefficients (β) for continuous outcomes and as odds ratios (ORs) with 95% confidence intervals for binary outcomes using the broom::tidy() and exp(coef()) functions. A composite hemolytic component was generated with the use of an equation generated from a previously published study. ^23^ The variables LDH, AST, total bilirubin, and reticulocyte count were first standardized (z-scores) and then subjected to principal component analysis using prcomp(). The first principal component— scaled so that higher values indicate greater hemolysis—was extracted and regressed on HbF/F-cell using a linear model adjusted for age and sex. For MSH patients, a modified hemolytic component lacking LDH was calculated.

In the MSH patients, to estimate the incremental effect per 1 pg increase in HbF/F-cell, regression models were fit separately within each cohort with a negative binomial regression models to account for overdispersion and covariates including age, sex, follow-up time, and baseline event rate. To facilitate interpretation, model estimates were transformed into percent protection per 1 pg increase in HbF/F-cell. Analyses were conducted using Python, with regression models implemented via the statsmodels package.

Pipelines were developed using tidyverse, broom, stats, and ggplot2 packages for transparency and reproducibility. Random-seed control (set.seed;2025) was applied before modeling and resampling steps. Four different ways of measuring HbF were tested using a statistical score (AIC) to elucidate clinical relevance. Scripts used for data cleaning, computation of MCH and HbF/F-cell, regression modeling, and figure generation are available upon reasonable request.

## Results

### Modelling MCH and HbF:HbS ratio

As MCH increased from 22 to 42 pg, HbF:HbS ratio decreased at all HbF levels from 5 to 30%. (**Fig. 1 A**) When %HbF is 30% and MCH is about normal (30 ± 5 pg) the HbF:HbS ratio exceeded 0.30 in all cells. For perspective, most untreated patients with HbS/F genotypes have a %HbF of 5-10%. According to our modelling, none of their RBCs reached the HbF:HbS threshold of 0.30, reflecting the customary severity of SCD. When the same range of HbF levels are compared at MCH of 25 pg and 35 pg, the lower MCH was associated with more cells reaching the putative protective HbF:HbS ratio of 0.30 (**Fig. 1 B**). These modelling results suggest that, by increasing the proportion of cells with a favorable intracellular stoichiometry of HbF to HbS, lower MCH enhances HbF-mediated protection independently of changes in %HbF. Conversely, increasing MCH with constant HbF increases HbS per cell, decreases the HbF:HbS ratio, and reduces the proportion of cells protected from HbS polymerization. At MCH 30 pg, ~6.9 pg HbF is required to maintain HbF:HbS = 0.30. Modelling suggested that to maintain the HbF:HbS ratio at 0.30, for every 2 pg increase in MCH, HbF must increase by ~0.46 pg. (**Fig. 1 C**)

**Figure 1.**
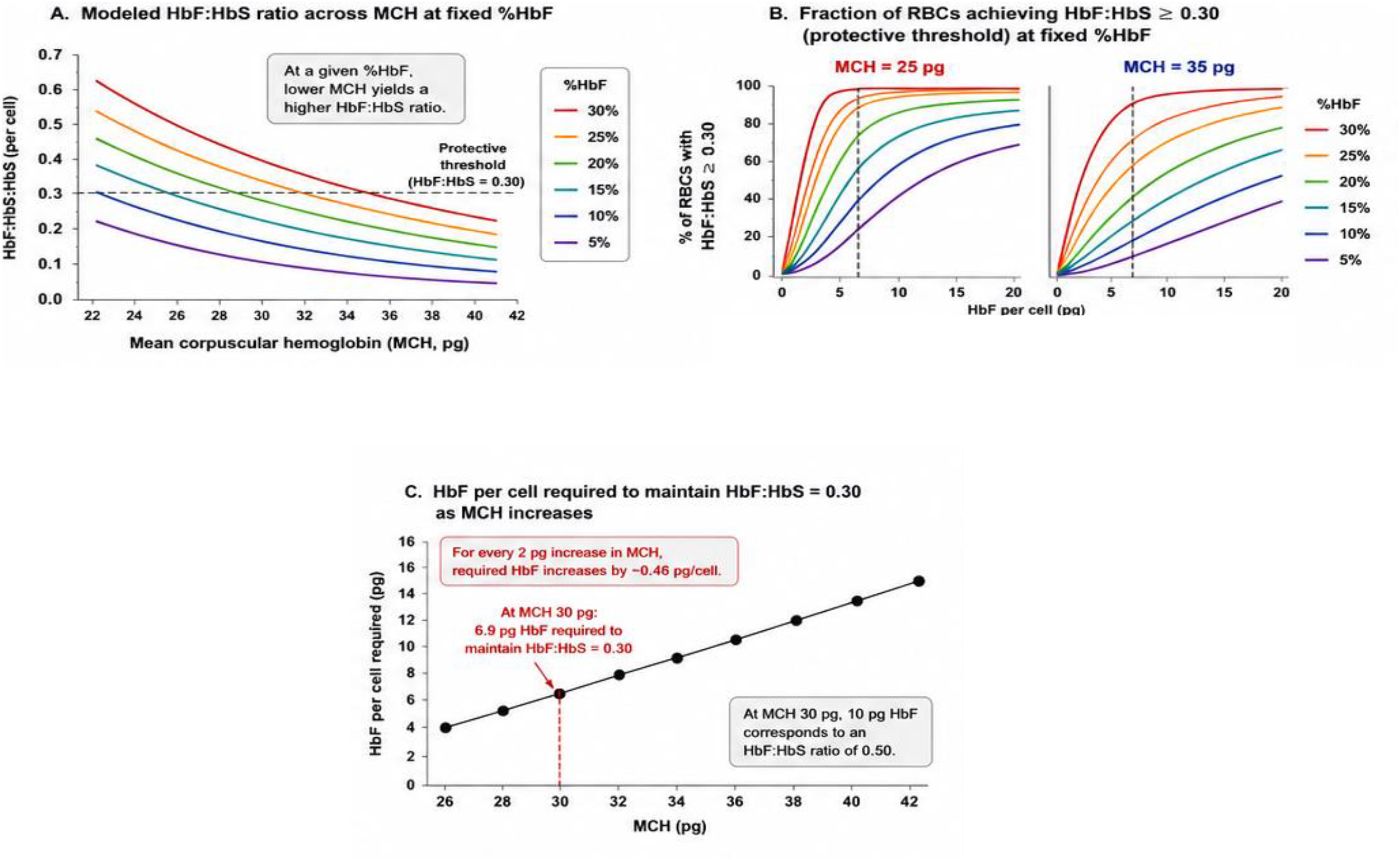

### Clinical data

Cohort characteristics are shown in **Table 1**. In the CSSCD cohort, the average number of VOEs was 7.52±15.1, 40.1% of patients had AVN, 8.8% had experienced stroke and 11.7% died. Of the 150 MSH cohort patients randomized to receive HU, average number of VOE was 42.4±64.2, average number of ACS was 1.10±2.4. In the BMC cohort followed 9.2±4.7 yrs, 76% were taking HU. Their combined annualized VOE + ACS rate was 18.1± 26.7 events.

### Application of Modeling to CSSCD, MSH and BMC Cohorts

The distribution of HbF/F-cells in the CSSCD cohort is shown in **Fig. S 1 A**. Distribution of HbF/F-cell by age deciles is shown in **Fig. S1 B**. Kruskal-Wallis test on HbF/F-cell across age deciles indicated that HbF/F-cell decreased significantly from the first to the third decile. Associations of HbF/F-cell and reticulocyte count, LDH, and hemoglobin concentration (adjusted for sex and age) in the CSSCD dataset are shown in **Figs. S 2 A, B, C and D**. We observed a positive association between hemoglobin concentration and HbF/F-cell (β = 0.22, p < 0.01) while negative relationships occurred between HbF/F-cell and reticulocyte count (β = −0.31, p < 0.01). No significant linear associations between LDH and HbF/F-cell was found, adjusting for sex and age. In **Fig. S 2 D**. HbF/F-cell was inversely correlated with the hemolytic component (β = −0.04, p < 0.01), indicating that higher intracellular HbF content is associated with less hemolysis.

HbF/F-cell and the association with VOEs, ACS, avascular necrosis (AVN), stroke, priapism, hemolytic component, and death in the CSSCD cohort are provided in **Table S 2** and **Fig. 2**. As HbF/F-cell increases, VOEs, ACS, stroke, AVN, and the likelihood of death decrease (**Fig. 2 A**) as does the hemolytic component modeled using linear regression, where the β coefficient (pg/dL) was −1.35 (−1.75 to −0.95). The estimated protection from VOEs and ACS per 1 pg increase in HbF/F-cell in the CSSCD was −5.8% (−6.7 to −4.9%) and −6.0% (−7.3 to −4.6%) respectively, while in the MSH cohort, VOEs fell −11.5% (−12.8 to −9.8%) and ACS, −13.5% (−15.2 to −11.6%). Protection was defined as the percent reduction in outcome rate per 1 pg increase in HbF/F-cell, calculated as (1 − rate ratio) × 100. We used CSSCD data to determine whether HbF/F-cell provides information beyond %HbF, by examining predicted VOE burden across tertiles of HbF/F-cell within strata of %HbF (**Fig. 2 B)**. Within strata of %HbF, HbF/F-cell remained strongly associated with predicted VOE burden, with clear separation across tertiles. Individuals with similar %HbF exhibited markedly different risks according to HbF/F-cell, demonstrating that HbF/F-cell provides meaningful discrimination beyond %HbF. These results indicate that HbF/F-cell captures biologically relevant variation in HbF distribution across red cells that is not reflected by %HbF alone. Across all outcomes, HbF/F-cell demonstrated stronger and more consistent associations with clinical severity than %HbF (**Fig. S 3**). In models including identical covariates, both %HbF and HbF/F-cell were associated with clinical outcomes; however, HbF/F-cell demonstrated consistently larger effect sizes across all endpoints. For example, HbF/F-cell was associated with a greater reduction in VOEs and stroke compared with %HbF per unit increase. Model fit statistics were similar between the two measures, with %HbF demonstrating slightly lower AIC values.

**Figure 2.**
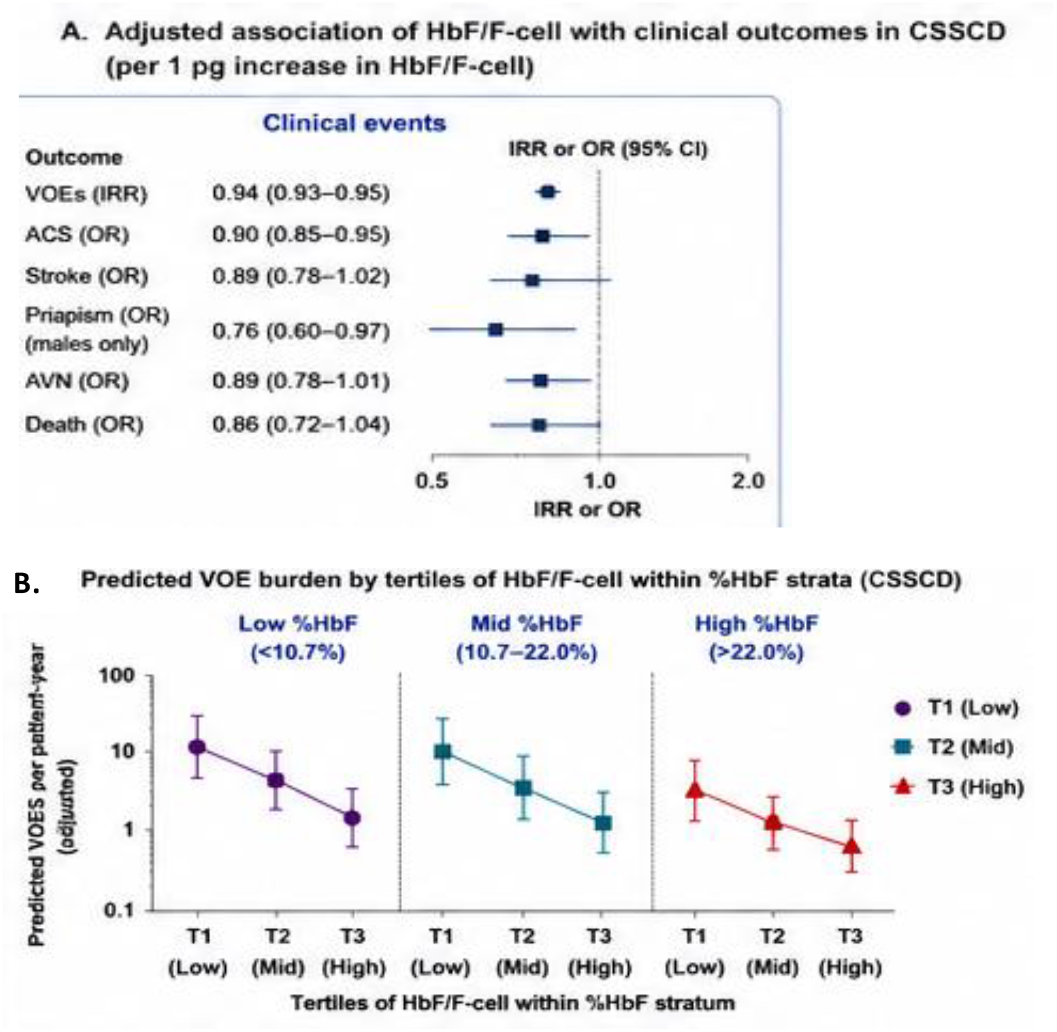

### ΔHbF/F-cell and clinical complications and hemolytic anemia in the MSH

**Fig. 3. A** displays the absolute change in HbF in placebo treated and HU treated MSH patients; **Fig. 3 B** shows associations of ΔHbF/F-cell with VOEs and hemolysis. With increasing ΔHbF/F-cell there is increasing reduction in VOE (Kruskal–Wallis p<0.001; p-trend <0.005) and a trend toward reduction in the hemolytic component.

In the BMC cohort, annualized VOEs and ACS were associated with HbF/F-cell; for each 1 pg increase in HbF/F-cell there was a 9.0% reduction in acute event rate (adjusted for age, sex, and HU usage). With each 1 pg increase in HbF/F-cell there was a 7.4% reduction in ACS (adjusted for age, sex, and HU use). Increased HbF/F-cell was also associated with a 4.0% decrease in hemolysis (adjusted for age, sex, and HU use) as estimated by the reticulocyte count.

**Figure 3.**
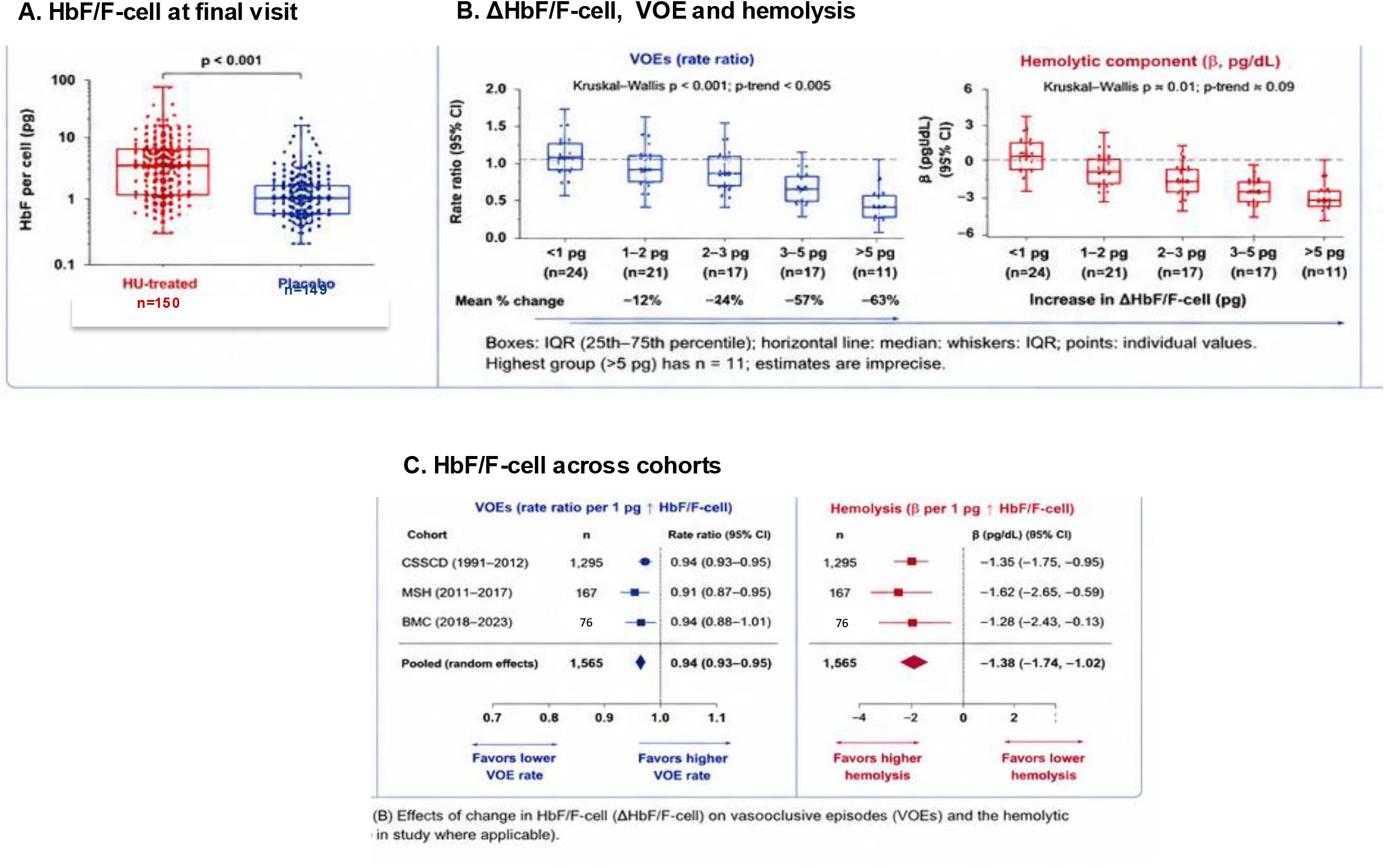

**Fig. 3. C** summarizes the results in the BMC, CSSCD, and MSH cohorts with congruent effects of HbF/F-cell on VOE and hemolysis.

## Discussion

Cellular HbF:HbS ratio reflects the relative proportion of anti-polymerizing HbF and polymerizing HbS. At a ratio of 0.30, the sickle RBC is protected from HbS polymer damage. HbF/F-cell quantifies the absolute amount of HbF/RBC and, in the context of MCH, approximates whether sufficient HbF is present to inhibit polymer formation. While correlated, HbF:HbS ratio and HbF/F-cell are not interchangeable. The HbF:HbS ratio differences between gene therapies and the best HU responders who can have %HbF approaching that achieved by gene therapy, are reflected clinically by a “functional “cure” in gene therapy patients while signs and symptoms of disease linger, albeit at reduced levels, in nearly all HU treated adults.^12, 17^

To develop a more clinically meaningful biomarker, we calculated mean HbF/F-cell and examined its association with sub-phenotypes of SCD. Among diverse and independent cohorts, both %HbF and HbF/F-cell were significantly associated with the improvement of multiple clinical and laboratory features of SCD. However, HbF/F-cell demonstrated stronger associations with clinical severity measures across cohorts. HbF/F-cell remained informative within three strata of %HbF, distinguishing patients at higher versus lower risk despite similar overall HbF levels. These findings suggest that HbF/F-cell is not just a surrogate for %HbF, but provides additional, physiologically relevant information reflecting the distribution of intracellular HbF concentrations and protection at the level of individual RBCs. While %HbF remains an important measure, especially at the population level, HbF/F-cell uncovers RBC**-**level protection, erythrocyte sickling biology, and, perhaps, explains the heterogeneity of clinical response even when %HbF levels are similar.

There are limitations to our analysis. Laboratory assays vary across sites and over time, potentially introducing measurement variability. The contemporary BMC cohort used to validate the older CSSCD and MSH is very small, but the results are congruent with these larger groups. A sizable contemporary cohort is being presently evaluated via a grant from the American Society of Hematology Research Collaborative. Iron deficiency reduces MCH and might be associated with reduced HbF synthesis, especially in patients taking HU.^24^. Patients with severe disease might be more intensively treated with HU or transfusions, complicating analyses. In all three cohorts, events can be under and over-reported. While we report associations and prognostic relationships, causality cannot be inferred from observational and retrospective data. Prognostic models may be prone to overfitting or limited transportability across independent cohorts. Importantly, our reported HbF/F-cell is an average. An F-cell assay that can quantitate the distribution of HbF concentrations among F-cells should be even more physiologically relevant than calculated HbF/F-cell.^25^ Functional assays estimating the RBC injury that is tempered by HbF, like variations of ektacytometry, imaging flow cytometry, and lensless digital holographic microscopy might also be useful.^26–29^ At present, most of these assays require sophisticated instrumentation and operator experience making them poorly suited for point-of-care deployment. A prospective study of HbF/F-cell as a predictive biomarker that replicates our findings would position this inexpensive, simple, and nearly universally available metric as an aide for clinical decision making and as an outcome variable in SCD clinical trials, especially HbF-inducing oral HbF inducers and gene therapies.

## Data Availability

All data produced in the present study are available upon reasonable request to the authors

## Acknowledgements

E.S.K. was supported by NIH/NHLBI 1UG3 HL143192 and HRSA U1EMC27864-08-00. and received research support from Novo Nordisk, Novartis, Bristol Myers Squibb, and United Therapeutics.

M.H.S. serves on the Steering Committee for the exa-cel trial of Vertex/CRISPR, is a member of the Scientific Advisory Board of Fulcrum Therapeutics, is a member of the iDMC of Cellarity, has served as a consultant for Beam and Editas Medicine, and is a member of the endpoint adjudication committees of sickle cell therapeutic trials of Noo Nordisk and Sanofi. E.S.K. is a consultant for Novo Nordisk.

## Supplemental figures

**Figure.**
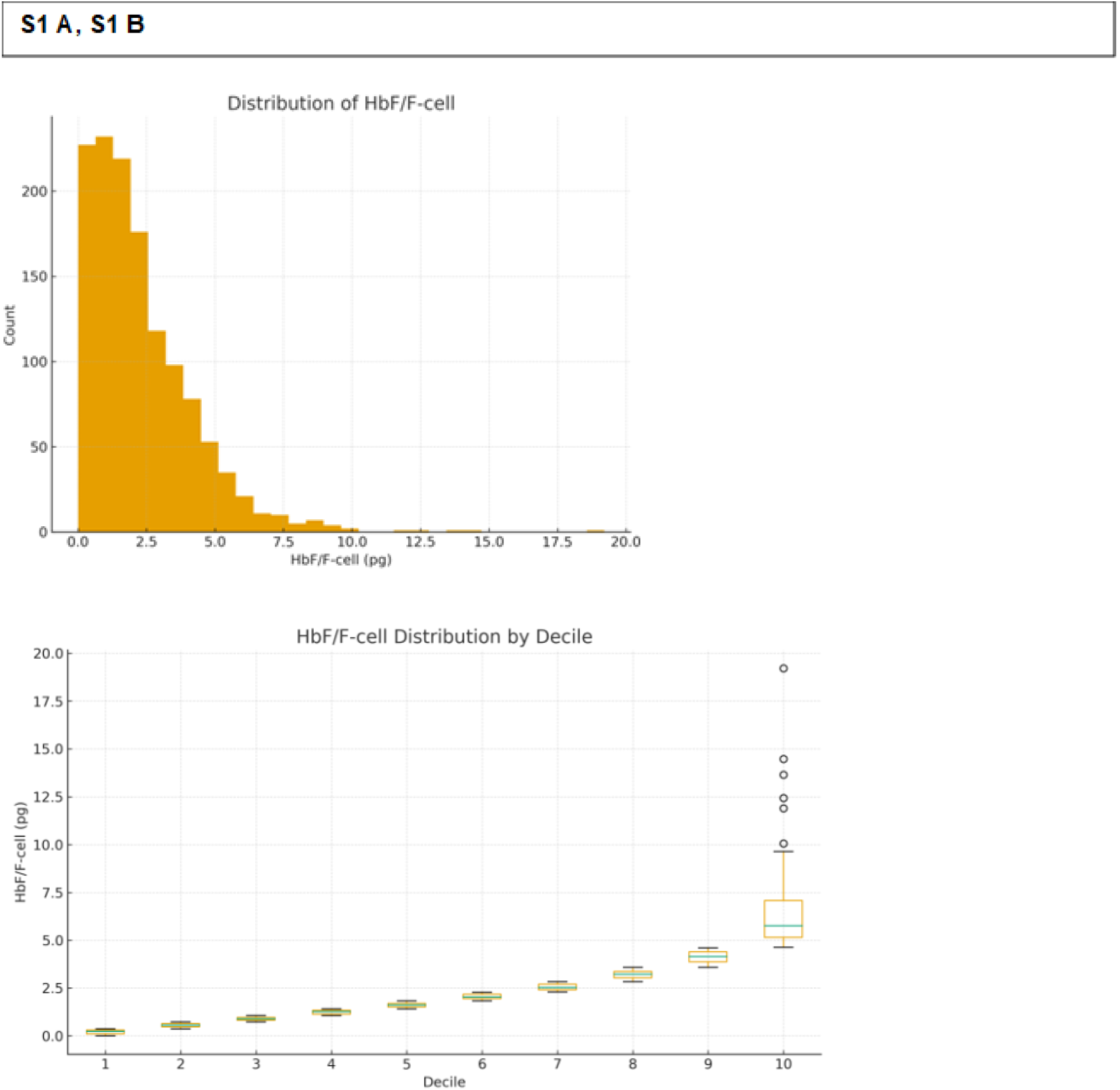
**S A**. Distribution of HbF/F-cells in the CSSCD cohort, the **S1B. Distribution of HbF/F-cell in CSSCD cohort**. Decile 1: N=131.0, Mean=0.210, SD=0.111; Decile 2: N=130.0, Mean=0.559, SD=0.100; Decile 3: N=130.0, Mean=0.893, SD=0.093; Decile 4: N=130.0, Mean=1.249, SD=0.105; Decile 5: N=130.0, Mean=1.613, SD=0.119; Decile 6: N=130.0, Mean=2.048, SD=0.139; Decile 7: N=130.0, Mean=2.550, SD=0.164; Decile 8: N=130.0, Mean=3.194, SD=0.208; Decile 9: N=130.0, Mean=4.135, SD=0.300; Decile 10: N=130.0, Mean=6.449, SD=2.102.

**Figure S2 A, B, C, D.**
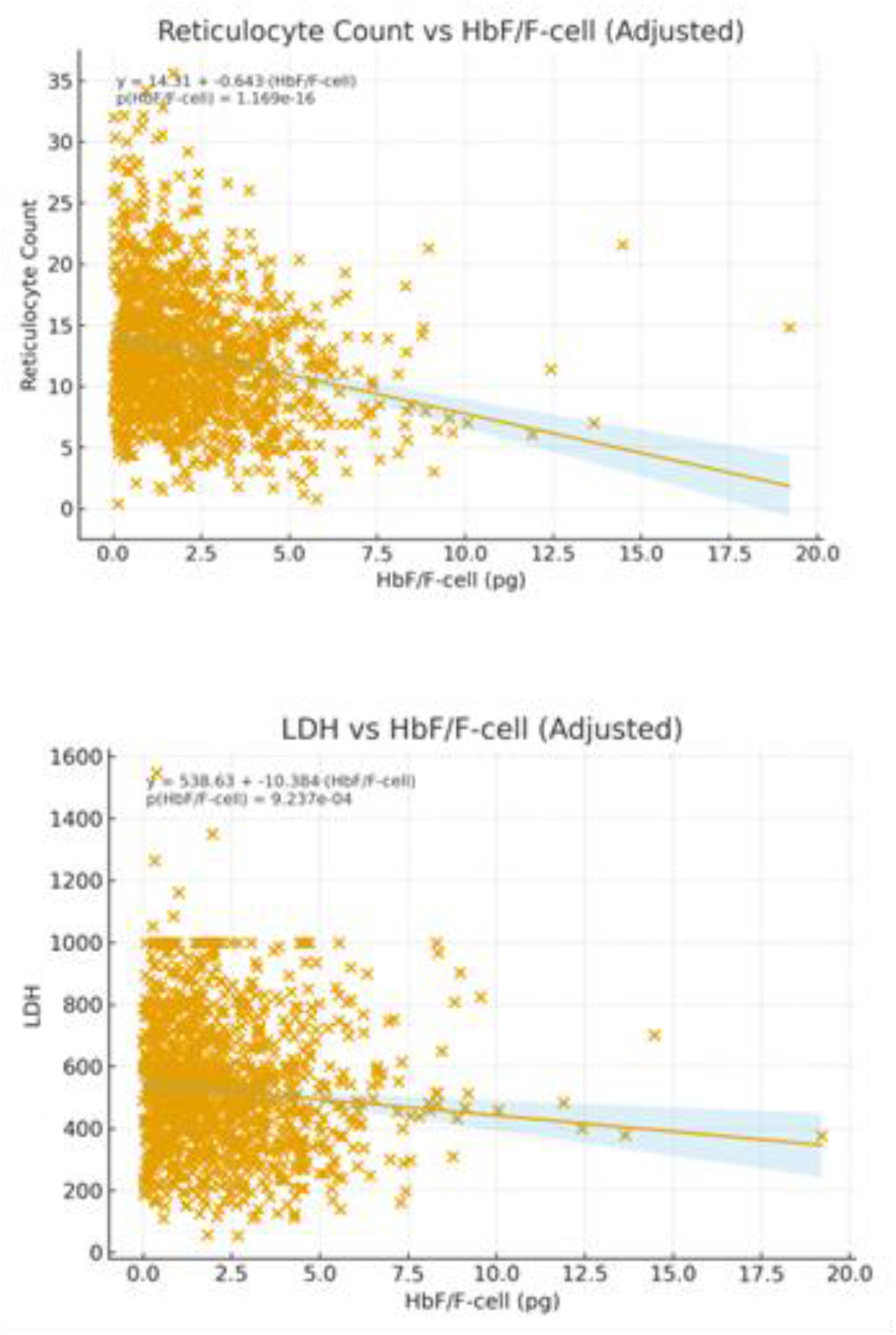
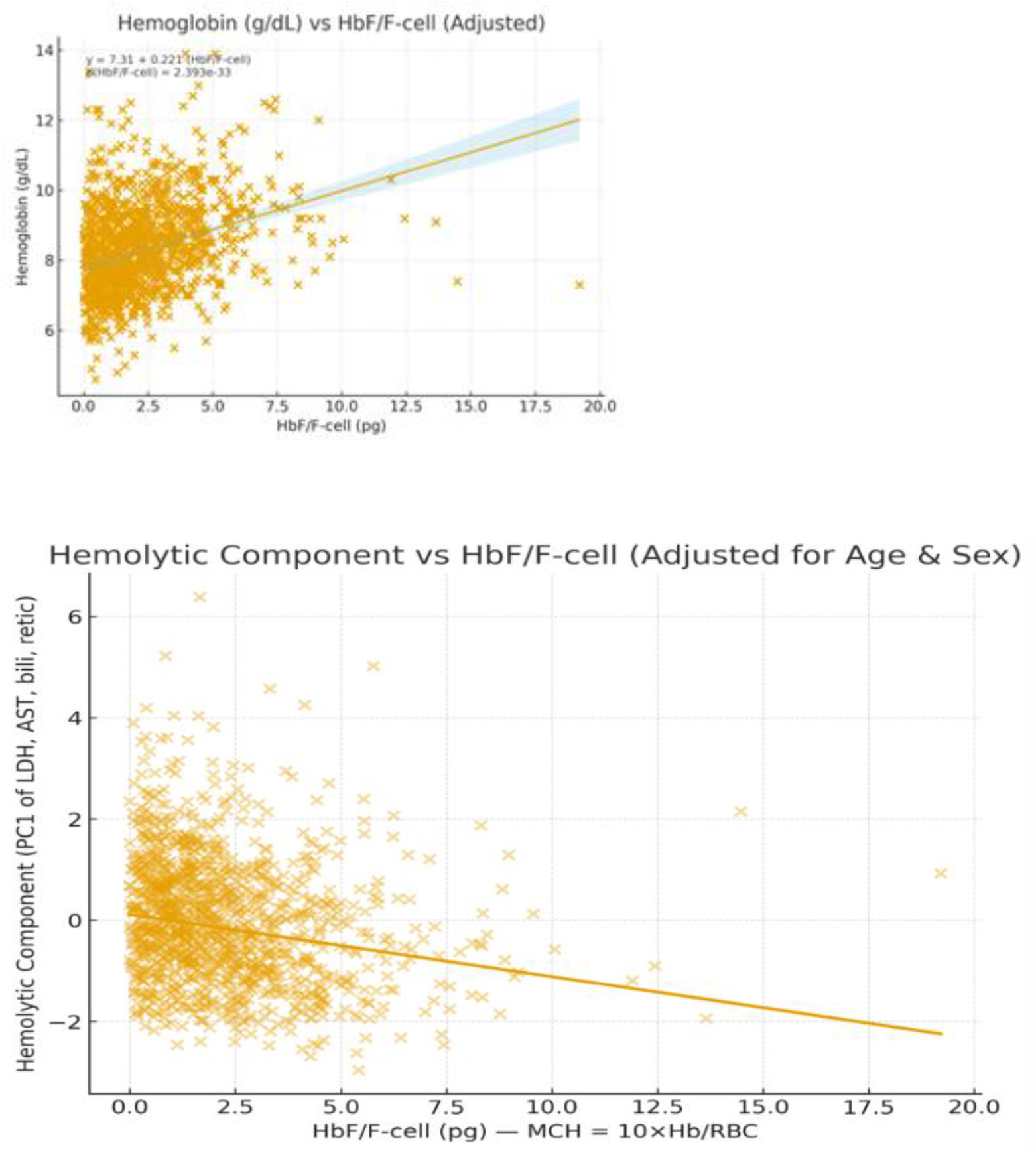
A. reticulocyte count, B. LDH, C. hemoglobin concentration, and D. hemolytic component. Reticulocyte count and hemolytic component decreased as HbF/F-cell increased. Hemoglobin increased as HbF/F-cell increased. The association with LDH was weak, though trended to w reduction in LDH with increased HbF/F-cell. Analysis was adjusted for age and sex.

**Figure S 3.**
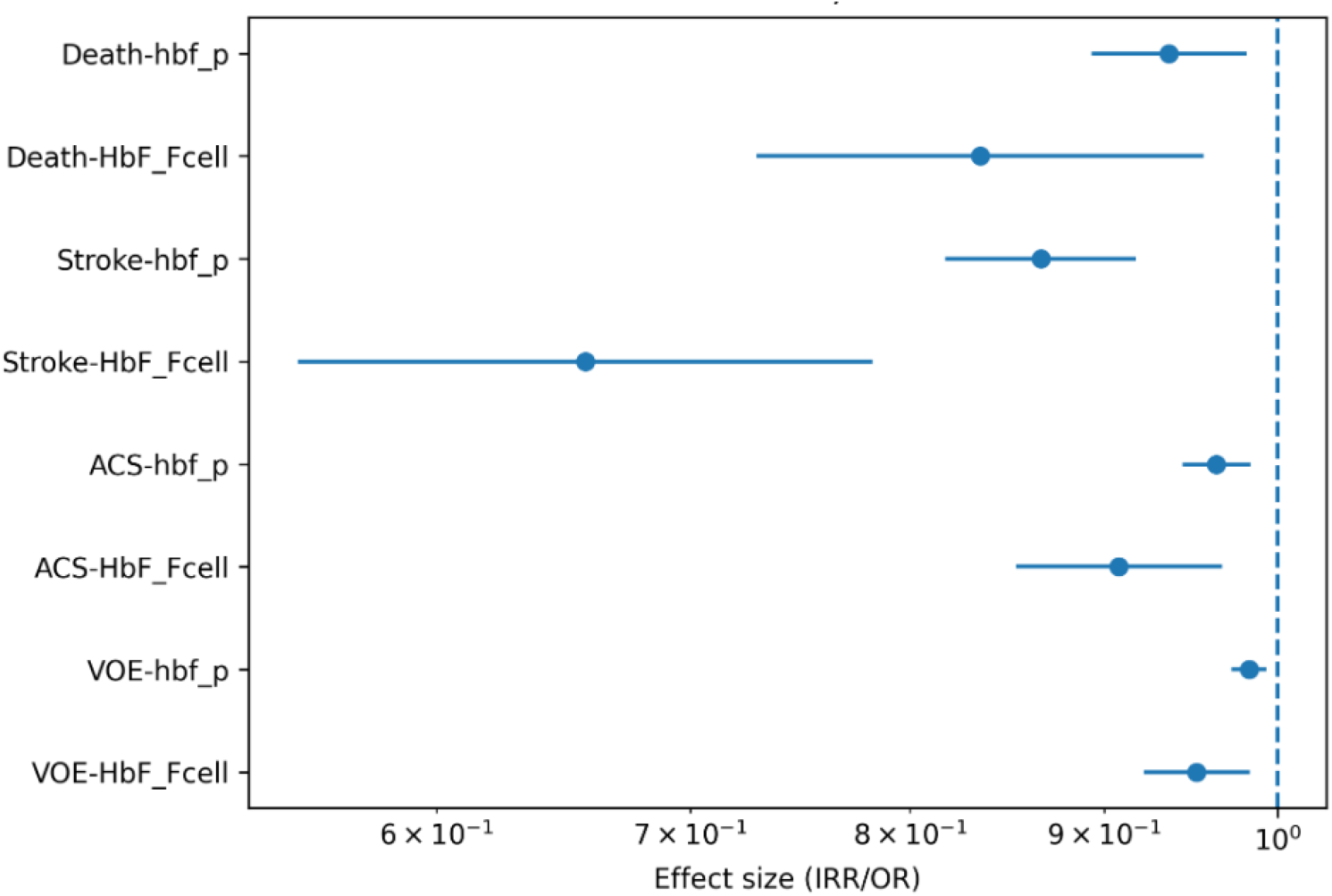
Comparative associations of HbF/F-cell and %HbF with clinical outcomes in the CSSCD. Associations of HbF/F-cell and %HbF with VOEs, ACS, stroke, and death are shown from multivariable regression models. Points represent effect estimates per unit increase in each biomarker (HbF/F-cell in pg and %HbF, and horizontal lines denote 95% confidence intervals. Effect estimates are expressed as incidence rate ratios (IRRs) for VOE and odds ratios (ORs) for binary outcomes, plotted on a logarithmic scale. HbF/F-cell shows consistently larger effect sizes compared with %HbF. Overall model fit, as assessed by Akaike Information Criterion (AIC), was similar between the two measures indicating that while both biomarkers are associated with clinical outcomes, HbF/F-cell has a stronger association with disease severity, consistent with its reflection of intracellular hemoglobin composition.

**Table S1.**
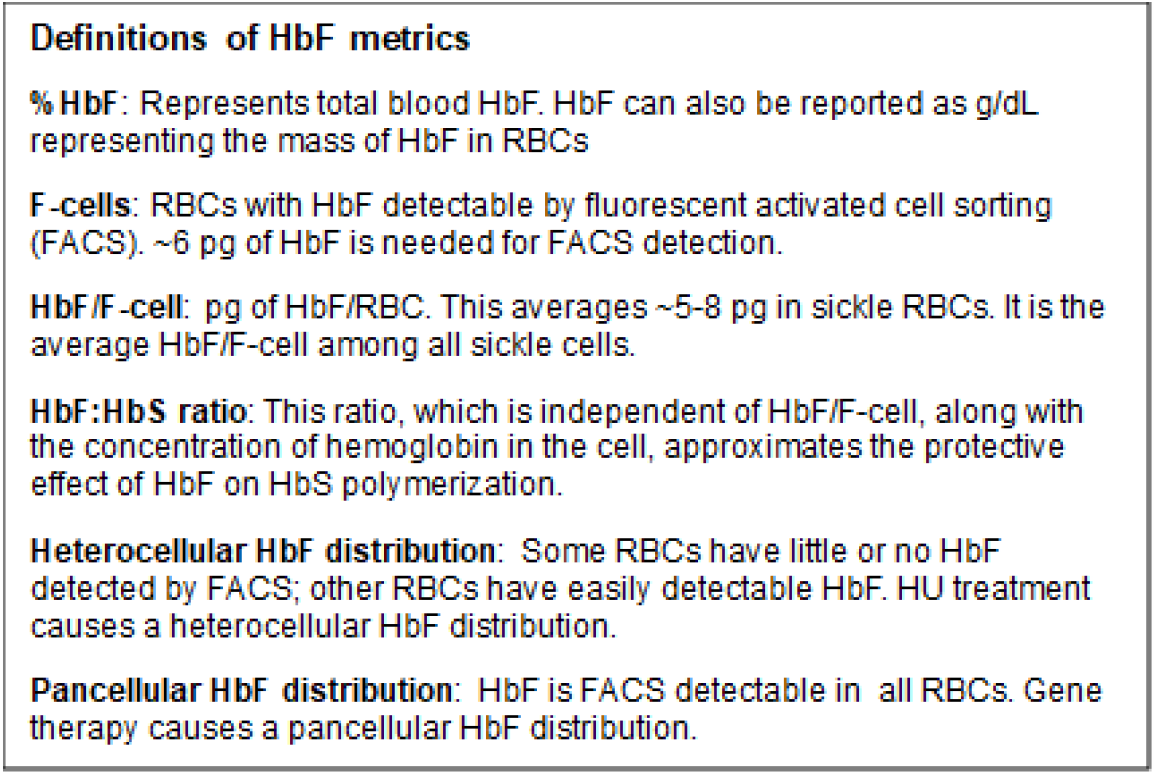
Hbf metrics.

**Table S2.** Association of HbF/F-cell with clinical outcomes in the CSSCD cohort. Associations between HbF/F-cell and clinical outcomes were evaluated in participants from CSSCD, restricted to individuals with HbS/F genotypes, aged ≥5 years. HbF/F-cell was calculated as MCH × (%HbF/100). VOEs were modeled as event counts using negative binomial regression with log(time in study) as an offset. Binary outcomes like ACS, stroke, priapism, AVN, and death were analyzed using logistic regression. The hemolytic component was analyzed using linear regression and defined as the first principal component of LDH, AST, bilirubin, and reticulocyte count; higher values reflect greater hemolysis. All models were adjusted for age, sex, and time in study, except for priapism. Effect estimates are presented per 1 pg increase in HbF/F-cell as incidence rate ratios (IRR) for VOE, odds ratios (OR) for binary outcomes, and regression coefficients (β) for the hemolytic component, with 95% confidence intervals (CI) and two-sided p-values. N are the number of cases for each outcome each outcome.

| Outcome | Model | N | Events | Effect | 95% CI | p-value |
| --- | --- | --- | --- | --- | --- | --- |
| VOE (count) | NegBin | 1277 | 9903 | IRR=0.952 | 0.914 to 0.992 | 0.0202 |
| Hemolytic component | Linear | 1215 | | $\beta=-0.121$ | -0.157 to -0.085 | 6.09E-11 |
| ACS | Logistic | 1277 | 898 | OR=0.908 | 0.853 to 0.967 | 0.00253 |
| Priapism (males) | Logistic | 600 | 130 | OR=0.931 | 0.825 to 1.052 | 0.254 |
| Stroke | Logistic | 1277 | 100 | OR=0.656 | 0.551 to 0.782 | 2.34E-06 |
| AVN | Logistic | 1277 | 503 | OR=0.906 | 0.850 to 0.967 | 0.00293 |
| Death | Logistic | 1277 | 127 | OR=0.829 | 0.723 to 0.952 | 0.0078 |

## Notes

### Author Declarations

Boston Medical Center and Boston University Medical Campus Institutional Review Board (IRB) - Study: H-35223. "Your amendment to update Internal Study Personnel has been reviewed and given administrative approval by the IRB because any added study personnel have been determined to meet institutional requirements. Please note that administrative approval of this amendment does not change the expiration or status check-in due date of this study. All investigators are required to meet the Institutional requirements for training. Amendment Description Adding Andrew Wilks, MD to the study team as a co-investigator."

## References

1. Sunshine HR, Hofrichter J, Eaton WA. Gelation of sickle cell hemoglobin in mixtures with normal adult and fetal hemoglobins. J Mol Biol. 1979;133:435–67.

2. Eaton WA, Hofrichter J. Sickle cell hemoglobin polymerization. Adv Protein Chem. 1990;40:63–279.

3. Akinsheye I, Alsultan A, Solovieff N, Ngo D, et al. Fetal hemoglobin in sickle cell anemia. Blood. 2011;118:19–27.

4. Platt OS, Brambilla DJ, Rosse WF, Milner PF, et al. Life expectancy and risk factors for early death. N Engl J Med. 1994;330:1639–44.

5. Maitra P, Caughey M, Robinson L, et al. Risk factors for mortality in adult patients with sickle cell disease: a meta-analysis of studies in North America and Europe. Haematologica. 2017;102:626–36.

6. Fitzhugh CD, Hsieh MM, Allen D, et al. Hydroxyurea-Increased fetal hemoglobin Is associated with less organ damage and longer survival in adults with sickle cell anemia. PLoS One. 2015;10:e0141706.

7. Boulassel M-R, Al-Badi A, Elshinawy M, et al. Hemoglobin F as a predictor of health related quality of life in children with sickle cell anemia. Qual Life Res. 2019;28(2): 473–9.

8. Sebastiani P, Steinberg MH. Fetal hemoglobin per erythrocyte (HbF/F-cell) after gene therapy for sickle cell anemia. Am J Hematol. 2023;98:E32–E34.

9. Boyer SH, Belding TK, Margolet L, Noyes AN. Fetal hemoglobin restriction to a few erythrocytes (F-cells) in normal human adults. Science. 1975;188:361–3.

10. Steinberg MH, Lu ZH, Barton FB, Terrin ML, Charache S, Dover GJ. Fetal hemoglobin in sickle cell anemia: determinants of response to hydroxyurea. Multicenter Study of Hydroxyurea. Blood. 1997;89:1078–88.

11. Ngo DA, Aygun B, Akinsheye I, Hankins JS, et al. Fetal haemoglobin levels and haematological characteristics of compound heterozygotes for haemoglobin S and deletional hereditary persistence of fetal haemoglobin. Br J Haematol. 2012;156:259–64.

12. Frangoul H, Locatelli F, Sharma A, Bhatia M, et al. Exagamglogene autotemcel for severe sickle cell disease. N Engl J Med. 2024;390:1649–62.

13. Gupta AO, Sharma A, Frangoul H, Kanter J, et al. Base editing of HBG1 and HBG2 promoters for sickle cell disease. N Engl J Med. 2026; DOI: 10.1056/NEJMoa2504835.

14. Hanna R, Frangoul H, Pineiro L, McKinney C, et al. CRISPR-Cas12a gene editing of HBG1 and HBG2 promoters to treat sickle cell disease. N Engl J Med. 2026;394:1281–91.

15. Charache S, Barton FB, Moore RD, Terrin ML, et al. Hydroxyurea and sickle cell anemia. Clinical utility of a myelosuppressive “switching” agent. The Multicenter Study of Hydroxyurea in Sickle Cell Anemia. Medicine (Baltimore). 1996;75:300–26.

16. Steinberg MH, Barton F, Castro O, Pegelow CH, et al. Effect of hydroxyurea on mortality and morbidity in adult sickle cell anemia: risks and benefits up to 9 years of treatment. JAMA. 2003;289:1645–51.

17. Steinberg MH, Kutlar A, Sebastiani P. Mean corpuscular hemoglobin modulates HbF/F- Cell and clinical response to gene therapy and hydroxyurea in sickle cell disease. Am J Hematol. 2025;100:1647–50.

18. Gaston M, Smith J, Gallagher D, Flournoy-Gill Z, et al. Recruitment in the Cooperative Study of Sickle Cell Disease (CSSCD). Control Clin Trials. 1987;8(4 Suppl):131S–140S.

19. Steinberg MH, Lu ZH, Barton FB, Terrin ML, et al. Fetal hemoglobin in sickle cell anemia: determinants of response to hydroxyurea. Multicenter Study of Hydroxyurea. Blood. 1997;89:1078–88.

20. Ohene-Frempong K, Weiner SJ, Sleeper LA, Miller ST, et al. Cerebrovascular accidents in sickle cell disease: rates and risk factors. Blood. 1998;91:288–94.

21. Vichinsky EP, Neumayr LD, Earles AN, Williams R, et al. Causes and outcomes of the acute chest syndrome in sickle cell disease. National Acute Chest Syndrome Study Group. N Engl J Med. 2000;342:1855–65.

22. Stephens AD, Angastiniotis M, Baysal E, Chan V, Davis B, Fucharoen S, Giordano PC, Hoyer JD, Mosca A, Wild B; International Council for The Standardisation of Haematology (ICSH).ICSH recommendations for the measurement of haemoglobin F. Int J Lab Hematol. 2012;34:14–20.

23. Gordeuk VR, Campbell A, Rana S, Nouraie M, et al. Relationship of erythropoietin, fetal hemoglobin, and hydroxyurea treatment to tricuspid regurgitation velocity in children with sickle cell disease. Blood. 2009;114:4639–44.

24. Habibi A, De Pierrefeu V, Bencheikh L, et al. Iron deficiency limited hydroxyurea— induced fetal hemoglobin and clinical efficacy in sickle cell disease. 2025; 67th Annual Meeting of Am Soc Hematol, Orlando FL, abstract 0299.

25. Hebert N, Rakotoson MG, Bodivit G, Audureau E, et al. Individual red blood cell fetal hemoglobin quantification allows to determine protective thresholds in sickle cell disease. Am J Hematol. 2020;951235–45.

26. Sadif A, Seu, H, Thaman E, et al. Automated oxygen gradient ektacytometry: a novel biomarker in sickle cell anemia. Front Physiol. 202;12:636609.

27. Rab MAE, Kanne CK, Boisson C, Bos J, et al. Oxygen gradient ektacytometry-derived biomarkers are associated with acute complications in sickle cell disease. Blood Adv. 2024;8:276–86.

28. Fertrin K, van Beers EJ, Samsel L, et al. Imaging flow cytometry documents incomplete resistance of human sickle F-cells to ex vivo hypoxia-induced sickling. Blood. 2014;124:658–60.

29. Kyeremah C, Paul AS, Haehn D, Duraisingh MT, et al. Enhanced detection of malaria infected red blood cells through phase driven classification. Scientific Reports. 2025;15(1):30733. doi:10.1038/s41598-025-12899-3.

